# Ultrasensitive detection of p24 in plasma samples from people with primary and chronic HIV-1 infection

**DOI:** 10.1101/2021.01.07.21249393

**Authors:** Caroline Passaes, Héloïse M. Delagreverie, Véronique Avettand-Fenoel, Annie David, Valérie Monceaux, Asma Essat, Michaela Müller-Trutwin, Darragh Duffy, Nathalie De Castro, Christine Rouzioux, Jean-Michel Molina, Laurence Meyer, Constance Delaugerre, Asier Saez-Cirion

**Affiliations:** Institut Pasteur, HIV Inflammation et Persistance, Paris, France; INSERM U944, Université de Paris, Paris, France; Laboratoire de Virologie, Hôpital Saint Louis, Assistance Publique-Hôpitaux de Paris, Paris, France; Assistance Publique-Hôpitaux de Paris, Service de Microbiologie Clinique, Hôpital Necker-Enfants Malades, Paris, France; Université de Paris, Faculté de Médecine, Paris, France; INSERM, U1016, CNRS, UMR8104, Institut Cochin, Paris, France; Centre de Recherche en Epidémiologie et Santé des Populations (CESP), Université Paris-Sud, Université Paris-Saclay, INSERM, Le KremLin-Bicêtre, France; Institut Pasteur, Translational Immunology Lab, Paris, France; INSERM U941, Université de Paris, Paris, France; Département de Maladies infectieuses, Hôpital Saint Louis, Assistance Publique-Hôpitaux de Paris, Paris, France

**Keywords:** HIV/AIDS, HIV p24 antigen, Gag p24, Simoa, ELISA, biomarker, plasma

## Abstract

HIV-1 Gag p24 has long been identified as an informative biomarker of HIV replication, disease progression and therapeutic efficacy, but the lower sensitivity of immunoassays in comparison to molecular tests and the interference with antibodies in chronic HIV infection limits its application for clinical monitoring. The development of ultrasensitive protein detection technologies may help overcoming these limitations. Here we evaluated whether immune-complex dissociation combined with ultrasensitive digital ELISA Simoa technology could be used to quantify p24 in plasma samples from people with HIV-1 infection. We found that, among different immune-complex dissociation methods, only acid-mediated dissociation was compatible with ultrasensitive p24 quantification by digital ELISA, strongly enhancing p24 detection at different stages of HIV-1 infection. We show that ultrasensitive p24 levels correlated positively with plasma HIV-RNA and HIV-DNA and negatively with CD4+ T cells in the samples from people with primary and chronic HIV-1 infection. In addition, p24 levels also correlated with plasma D-dimers and IFNα levels. P24 levels sharply decreased to undetectable levels after initiation of combined antiretroviral treatment (cART). However, we identified a group of people who, 48 weeks after cART initiation, had detectable p24 levels despite having undetectable viral loads. These people had different virologic and immunologic baseline characteristics when compared with people who had undetectable p24 after cART. These results demonstrate that ultrasensitive p24 analysis provides an efficient and robust mean to monitor p24 antigen in plasma samples from people with HIV-1 infection, including during antiretroviral treatment, and may provide complementary information to other commonly used biomarkers.

**Importance:** The introduction of combined antiretroviral treatment has transformed HIV-1 infection in a manageable condition. In this context, there is a need for additional biomarkers to monitor HIV-1 residual disease or the outcome of new interventions, such as in the case of HIV cure strategies. The p24 antigen has a long half-live outside viral particles and it is therefore a very promising marker to monitor episodes of viral replication or transient activation of the viral reservoir. However, the formation of immune-complexes with anti-p24 antibodies difficult its quantification beyond acute HIV-1 infection. We show here that, upon immune-complex dissociation, new technologies allow the ultrasensitive p24 quantification in plasma samples throughout HIV-1 infection, at levels close to that of viral RNA and DNA determinations. Our results further indicate that ultrasensitive p24 quantification may have added value when used in combination with other classic clinical biomarkers.

## Introduction

Human Immunodeficiency Virus (HIV) capsid p24 is a 24-25kDa protein encoded by the *gag* gene and corresponds to the most abundant viral antigen. Detection of p24 is considered essential for the diagnosis of HIV infection, especially in infants born to women with HIV, individuals under pre-exposure prophylaxis and newly infected individuals (1). Fourth generation antibody-antigen assays detecting p24 in addition to anti-HIV antibodies are currently recommended for HIV testing (2), as they reduce the time between infection and HIV diagnosis allowing people with HIV (PWH) to start antiretroviral treatment as early as possible (3). Prior to the development of nucleic acid tests (NAT), p24 was used as a surrogate marker of HIV replication, disease progression and therapeutic efficacy in the clinical monitoring of PWH (4-10). At primary infection, p24 levels are increased and were shown to correlate with HIV-RNA (4, 11-16). At chronic HIV infection, PWH positive for p24 antigen showed a higher risk of developing AIDS than those who remain antigen negative (12, 17, 18). The quantification of plasma p24 has shown superiority to RNA viral determination in anticipating decline in CD4+ T cell counts in some studies (5, 18) and has shown to better correlate with CD8+ T cell activation in another (6). Moreover, p24 is sometimes detected dissociated from viral particles (i.e. in the absence of viral RNA), which might provide a more extensive estimation of the infection burden in the organism (6, 19).

However, although HIV-1 p24 is recognized as an informative biomarker, its detection by classical enzyme-linked immunosorbent assays (ELISAs) is less sensitive than HIV nucleic acid tests, in part due to the small sample volume that can be tested. In addition, after seroconversion, the development of anti-HIV humoral responses make the detection of p24 inaccurate by immunoassays due to the formation of antigen-antibody immune-complexes (1). The development of Single-molecule array (Simoa) technology represents an important recent advance in ultrasensitive protein detection, reaching detection at femtomolar concentrations (20, 21). With this technology, single molecule HIV-1 Gag p24 detection is 1,000-fold more sensitive than current available ELISA tests. We have previously tested the sensitivity of this assay in HIV-infected cells and in culture supernatants. We showed that viral proteins produced by a single infected cell could be detected by an ultrasensitive p24 assay, which opens new opportunities for the study of HIV pathogenesis (22). To date, the applicability of an ultrasensitive HIV p24 assay in patients’ samples was demonstrated only in serum from HIV-infected individuals during acute infection positive for HIV-RNA, but non-reactive for anti-HIV antibodies (23). More recently, the single molecule digital p24 assay showed enhanced sensitivity when compared to the currently approved fourth-generation antigen/antibody combination and stand-alone p24 antigen assays in a panel of serially diluted RNA-positive/antibody-negative plasma-derived culture supernatants (24). The applicability and relevance of ultrasensitive HIV p24 detection in serum/plasma samples from chronically infected individuals remains unknown.

We show here that, upon immune-complex dissociation (ICD) with an approach compatible with the single molecule assay, p24 can be detected at ultrasensitive levels in plasma from most individuals during primary and chronic HIV-1 infection. The levels of p24 dropped sharply upon antiretroviral treatment initiation but remained detectable in some individuals despite undetectable RNA viral load. The increased sensitivity of this technique could bring new insights for the predictive value of p24 as an informative biomarker of viral replication, inflammation and treatment efficacy in people with primary and chronic HIV-1 infection.

## Results

### Acid dissociation to disrupt antigen-antibody immune-complexes is compatible with ultrasensitive p24 quantification

Immune-complex dissociation is required to increase the sensitivity of p24 antigen assays during chronic HIV-1 infection. We therefore assessed whether common methods for ICD are compatible with the digital ELISA technology. We tested (i) heat, (ii) heat with SDS and DTPA, and (iii) acid dissociation with Glycine-HCl pH 1.8 or 2.5. Heat treatment denatures antibodies, but high and non-specific loss of antigen and protein coagulation due to this procedure was reported (25). The treatment with a solution containing SDS and DTPA for diluting samples prior to heat treatment aimed at reducing these problems. SDS reduces protein coagulation by giving them a negative charge and DTPA chelates iron and other ions, preventing further interference in the p24 assay (26).

We first evaluated the impact of these different methods on the ultrasensitive p24 standard curve. Based on the average enzyme per bead (AEB) and the shape of the standard curves obtained after different treatments for ICD, we observed that heat alone or in combination with SDS+DTPA importantly interferred with p24 determination, flattening the standard curves and increasing the background signal in the absence of p24 (Figures 1A and 1B). Higher AEB values were also observed when testing only the SDS+DTPA solution in the absence of p24 protein. The other reagents used for sample dilutions and virus inactivation did not shown any impact (Figure 1C). We next evaluated the effect of heat-mediated with SDS and DTPA and acid dissociation methods in plasma from HIV-1 negative donors. We confirmed that heat-mediated with SDS+DTPA treatment, but not acid dissociation, increases the background signal for p24 detection in plasma samples (Figure 1D). These results suggest that heat-mediated dissociation, with or without SDS+DTPA, is not compatible with the Simoa technology and may overestimate p24 levels in clinical samples. It is possible that heating itself alters the p24 conformation or that heating-mediated protein coagulation promotes non-specific binding of assay antibodies. Treatment with SDS/DTPA did not prevent these problems, conversely it contributed to an increase of non-specific signal.

**Figure 1.**
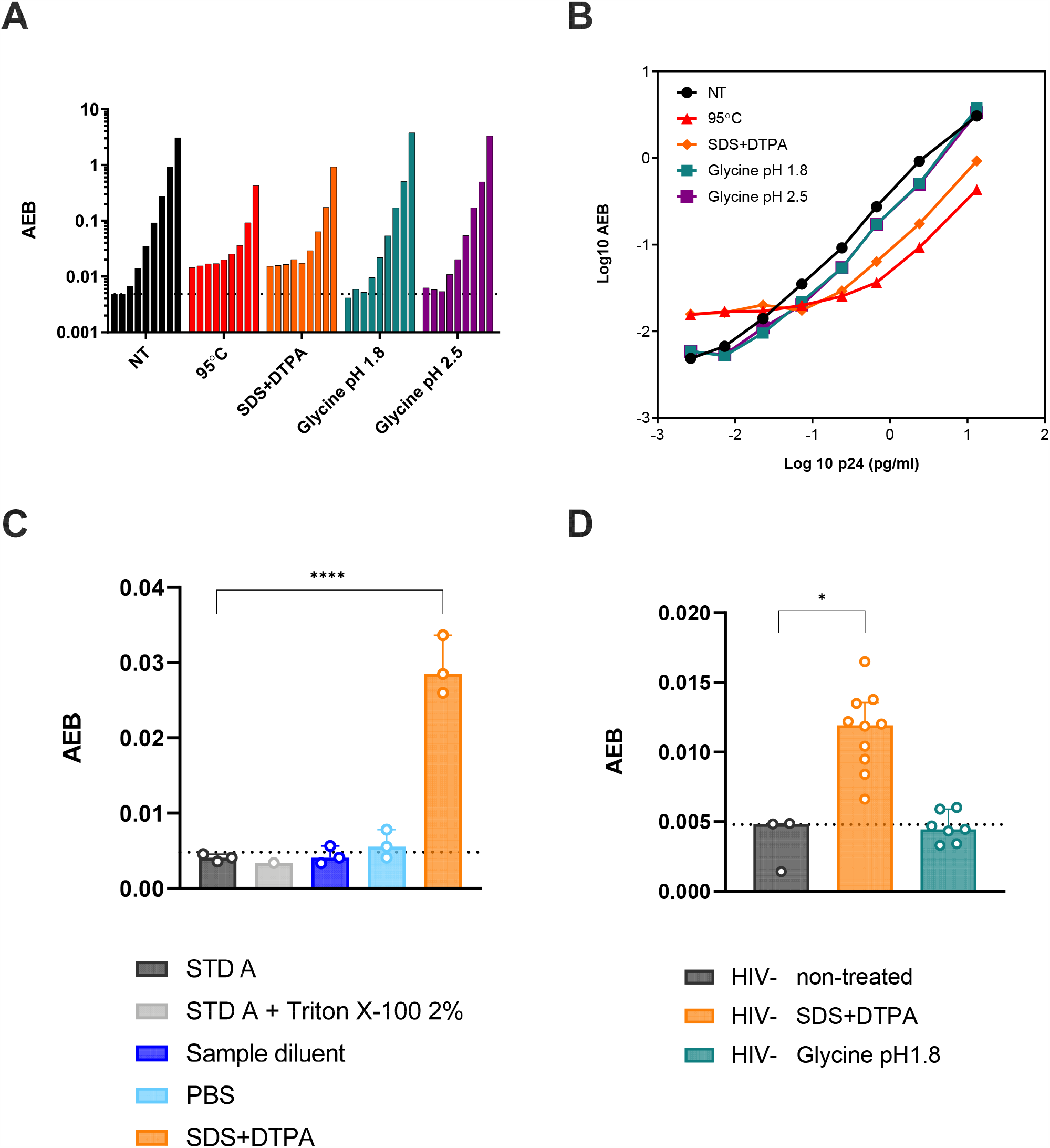
Acid dissociation to disrupt antigen-antibodies immune-complexes is compatible with ultrasensitive p24 quantification. **A)** Bars denote the average enzymes per beads (AEB) for: 0; 0.01; 0.02; 0.07; 0.24; 0.67; 2.39 and 13.17 pg/mL of p24 standards tested in different experimental conditions for immune-complex dissociation, as follows: non-treated (black), heat-mediated (red), heat-mediated with SDS and DTPA (orange) and acid dissociation with Glycine-HCl pH 1.8 (blue) or pH 2.5 (violet). **B)** Four-parameter logistic regression curves obtained from different experimental conditions for immune-complex dissociation, as follows: non-treated (black), heat-mediated (red), heat-mediated with SDS and DTPA (orange) and acid dissociation with Glycine-HCl pH 1.8 (blue) or pH 2.5 (violet). **C)** AEB values obtained for Standard A (p24 = 0 pg/mL, negative control) (dark gray), Standard A + Triton X-100 2% (light gray), Sample diluent manufactured by Quanterix (dark blue), PBS 1x (light blue) and the solution of 7mM SDS + 1.5 mM DTPA pH 7.2 (orange). Dashed line indicates the median value obtained for Standard A condition. **D)** AEB values obtained from plasma samples from HIV-1 negative donors non-treated (gray), after heat-mediated dissociation with SDS and DTPA (orange) and after acid dissociation with Glycine-HCl pH 1.8 (green). Dashed line indicates the median value obtained for non-treated condition.

In contrast, the curves obtained after acid-mediated dissociation with Glycine-HCl displayed the same profile than non-treated samples (Figures 1A, 1B and 1D). AEB values were comparable and standard curves obtained with non-treated samples and samples undergoing acid-mediated ICD presented equal distributions (Figures 1A and 1B). Therefore, the acid-mediated ICD methods described here can be used for p24 ultrasensitive quantification by digital ELISA, while other classical methods previously validated for ICD in classical ELISA assays are not compatible with this new technology. No differences were observed between Glycine pH 1.8 and pH 2.5, and we kept using the Glycine pH 1.8 protocol for further analyses.

### Acid-mediated ICD combined with _us_p24 assay enhances p24 quantification in plasma samples of people with HIV

After defining the best experimental conditions to quantify the levels of p24 in the plasma, we evaluated the sensitivity of Simoa p24 detection in plasma samples from PWH. We first determined a cutoff to best discriminate between positive and negative samples by analyzing plasma from HIV negative donors (n=15). Based on the results obtained, we determined 24 fg/mL (calculated as 2.5 standard deviations from the mean of p24 signal in plasma from HIV negative donors) as the cutoff to consider an ICD-treated plasma sample as positive for p24 in our experimental conditions (Figure 2A), and we used this cutoff in our analyses. This value is higher than the lower limit of quantification (10 fg/mL) and the lower limit of detection (LoD, 3 fg/mL) proposed for the assay, likely due to interference by some blood components in protein quantification.

**Figure 2.**
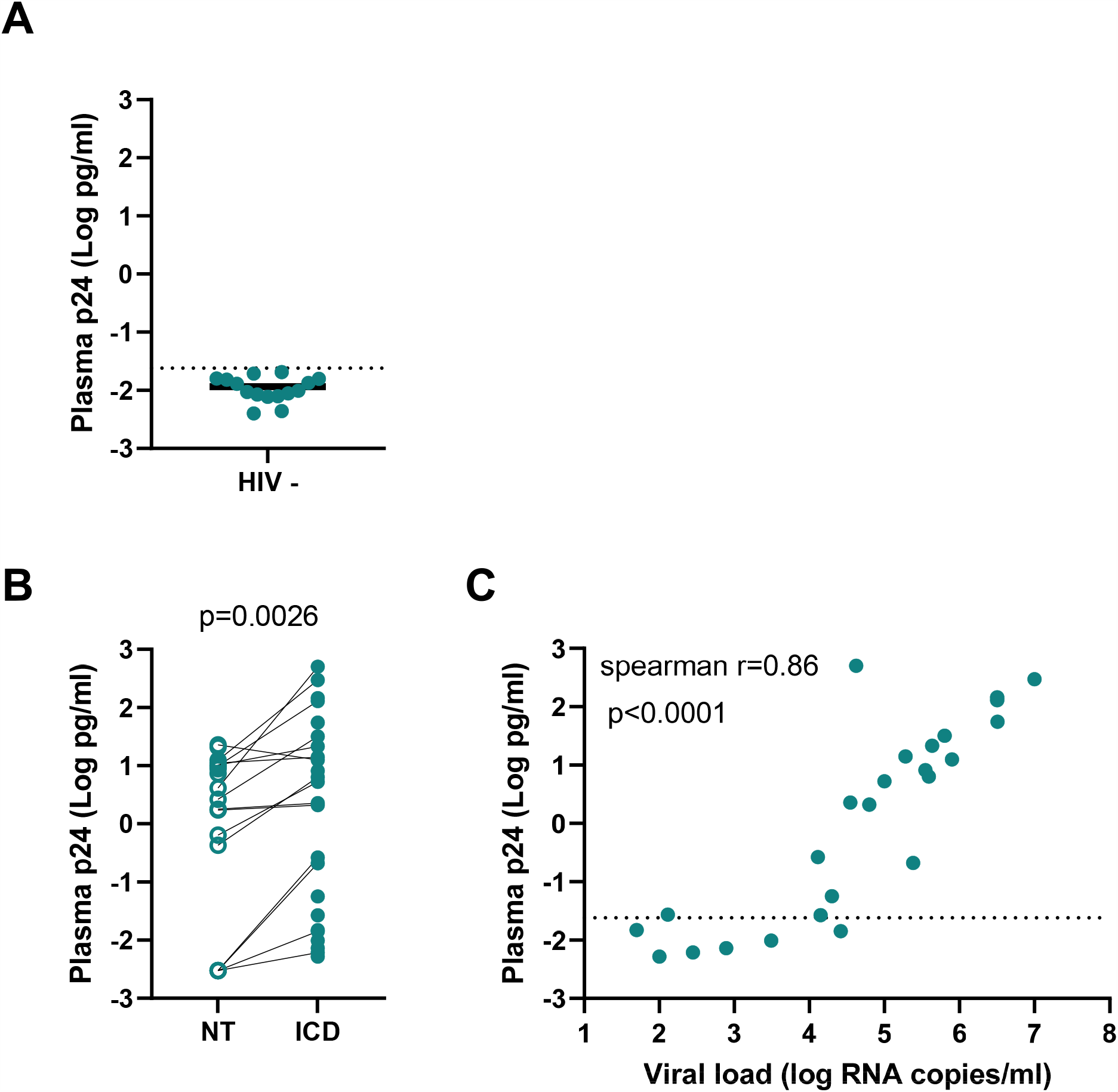
Sensitivity of ultrasensitive p24 quantification in plasma samples. **A)** p24 signal in plasma samples of HIV-1 negative donors. Samples received acid dissociation treatment with Glycine-HCl pH 1.8. Median is indicated by black line. Dashed line denotes the cutoff value of 0.024 pg/mL calculated as 2.5 standard deviations from the mean of p24 signal in the plasma from HIV negative donors. **B)** p24 levels detected in plasma samples from HIV-1 infected individuals non-treated for immune-complex dissociation (open symbols) and after acid dissociation with Glycine-HCl pH 1.8 (green dots). **C)** Relationship between positive p24 and HIV-RNA in plasma samples from HIV-1 infected individuals. Correlation was calculated using a nonparametric Spearman test.

We next evaluated the levels of p24 in plasma samples from 25 PWH displaying a wide range of detectable viremia ranging from 1.7 to 7 HIV-1 RNA Log copies/mL. The treatment of plasma samples with Glycine pH 1.8 for ICD increased the p24 levels detected compared to non-treated samples (p=0.0026) (Figure 2B). After ICD, 19/25 samples had p24 levels above our experimental cutoff. While p24 was often detected in samples with VL >10^4^ copies/mL (18/19 positive samples), the sensitivity dropped sharply for samples with lower VL (1/6 positive samples). Indeed, we found a strong correlation between p24 values and the corresponding plasma viral loads (Figure 2C; r= 0.86, p<0.0001). The correlation was still statistically significant when we considered only the samples with p24 values above the cutoff (r=0.76, p<0.0001). This correlation with viral RNA was not observed when we used the p24 values obtained in samples not treated by ICD (r=0.41, p=0.12). These analyses showed that immune-complex dissociation enhanced the ultrasensitive quantification of p24 in the plasma of PWH.

### Ultrasensitive p24 detection in a cohort of PWH during primary HIV-1 infection

To validate our aproach, we analyzed the ultrasensitive p24 quantification in 92 plasma samples from a subcohort of people with primary HIV-1 infection from the ANRS CO6 PRIMO cohort (27). Samples from 92 individuals were analyzed displaying highly variable plasma RNA viral loads (Figure 3A, 48,263 copies/mL [24 – 211,360,000]). The median time since estimated HIV acquisition was 52 days (range 20-183 days). We detected p24 values above the experimental cutoff in 72 of the 92 samples (Figure 3A). There was a wide range of p24 antigen concentrations (median 1.013 pg/mL; IQR [0.027 – 191.4]). As before, a strong correlation was observed between viral RNA and ultrasensitive p24 VL in the plasma samples analyzed (Figure 3B; r=0.7791, p<0.0001). We also found a strong positive correlation between _us_p24 levels and the cell-associated HIV-DNA (Figure 3C, r=0.64, p<0.0001). These correlations were still statistically significant when we considered only the samples with p24 values above the cutoff (r=0.69, p<0.0001 and r=0.61, p<0.0001 for HIV-RNA levels and HIV-DNA levels respectively). A weak negative correlation was observed between _us_p24 levels and the CD4+ T cell counts corresponding to these samples (Figure 3D; r=-0.22, p=0.03), although only a tendency was found with samples with above cutoff-p24 levels (r=-0.22, p=0.07).

**Figure 3.**
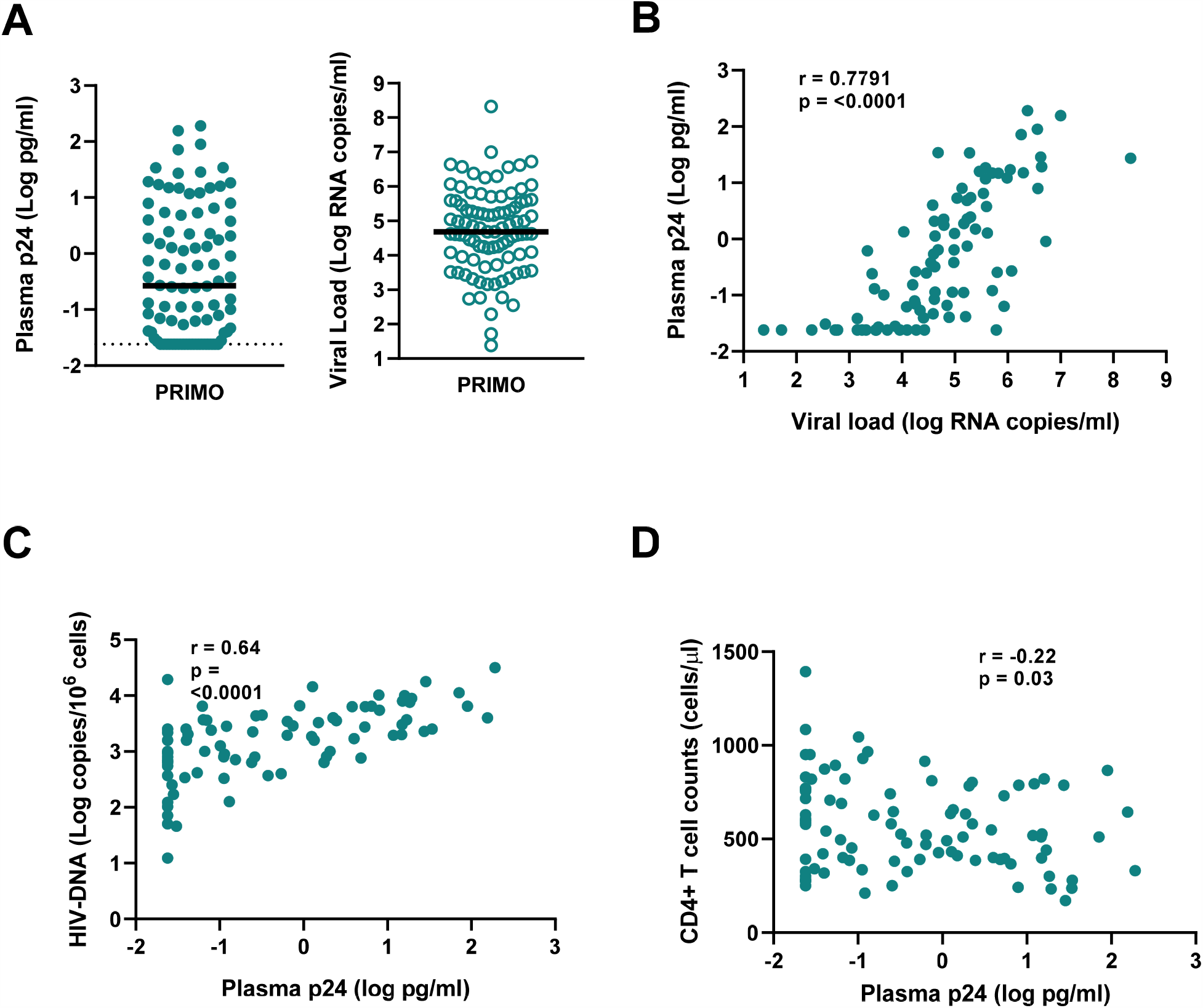
Ultrasensitive p24 detection in a cohort of acutely HIV infected individuals. **A)** *Left panel:* p24 levels in plasma samples from 92 individuals acutely infected with HIV-1 (ANRS PRIMO Cohort). Acid dissociation with Glycine-HCl pH 1.8 was used to disrupt immune-complexes. Samples below the limit of quantification were given an arbitrary value of 0.024 pg/mL based on the established cutoff. *Right panel:* HIV-RNA levels in plasma samples from the same 92 individuals acutely infected with HIV-1. Median is indicated by black line. Relationship between **B)** p24 and HIV-RNA, **C)** p24 and HIV-DNA and **D)** p24 and CD4+ T cell counts in plasma samples from individuals acutely infected with HIV-1. Correlations were calculated using a nonparametric Spearman test.

These data showed that _us_p24 can be quantified after ICD in plasma samples from PWH in different stages of primary infection. Moreover, the p24 levels were associated with other classic biomarkers of HIV-1 infection.

### Ultrasensitive p24 quantification in PWH at chronic HIV-1 infection prior to and after cART initiation

ICD should enable the quantification of p24 during chronic infection despite the presence of anti-p24 antibodies. We thus quantified _us_p24 in plasma samples from 137 volunteers with chronic HIV-1 infection (viral loads are depicted in Figure 4A) who were enrolled in the ANRS REFLATE TB trial and initiated cART treatment. Before cART (W0), p24 values above cutoff were detected in 98 out of 137 samples (Figure 4A). There was again a statistically significant correlation between _us_p24 and the RNAviral load in the plasmas analyzed (Figure 4B, r=0.42, p<0.0001). We also found a positive correlation between _us_p24 levels and CD4+ T cell associated HIV-DNA levels (Figure 4C, r=0.32, p=0.0004) and a tendency for a weak negative correlation with CD4+ T cell counts (Figure 4D, r=-0.15, p=0.07).

**Figure 4.**
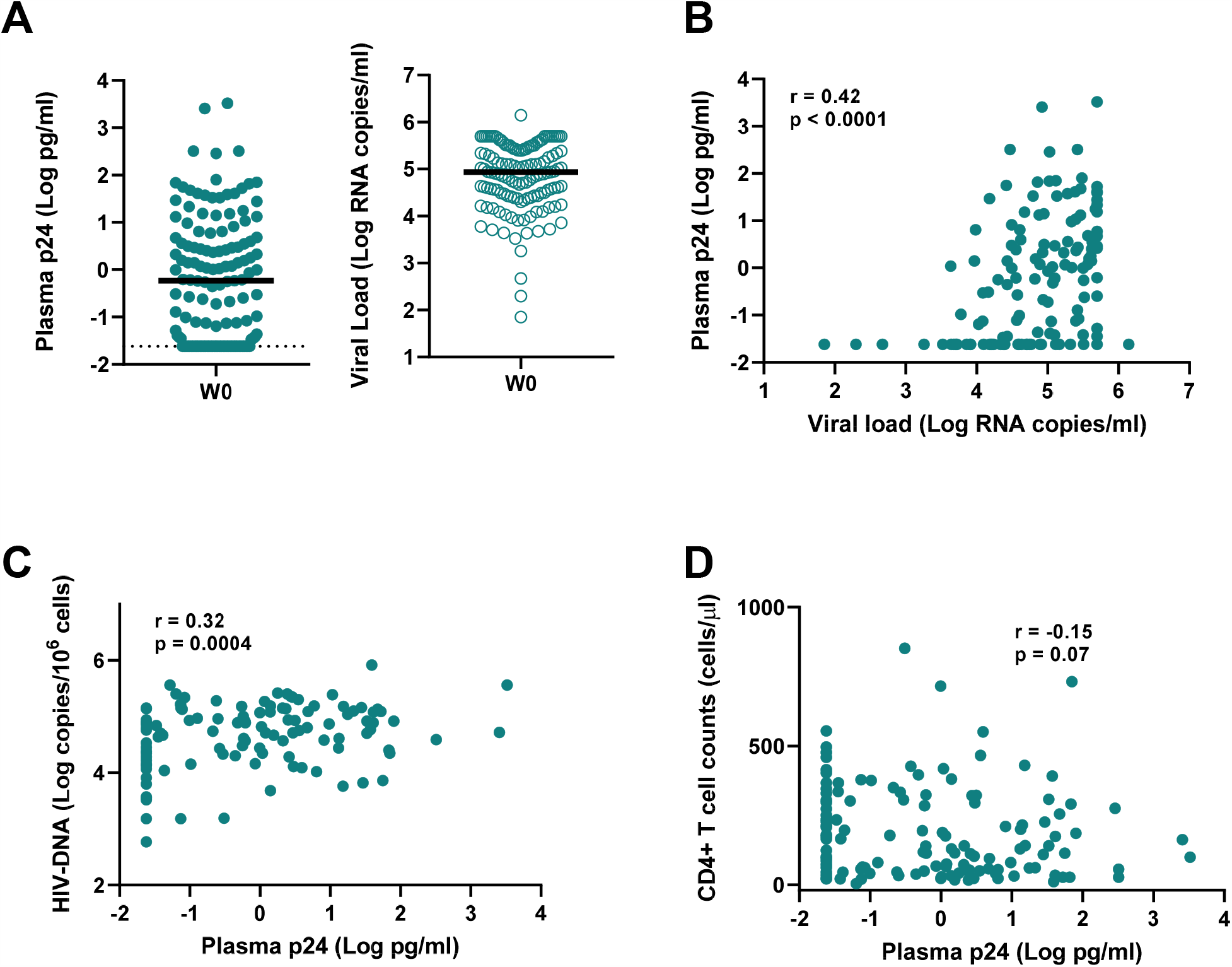
Ultrasensitive p24 detection in a cohort of chronically HIV infected individuals. *Left panel:* p24 levels in plasma samples from 137 individuals chronically infected with HIV-1 (ANRS 12180 Reflate TB trial). Acid dissociation with Glycine-HCl pH 1.8 was used to disrupt immune-complexes. Samples below the limit of quantification were given an arbitrary value of 0.024 pg/mL based on the established cutoff. *Right panel:* HIV-RNA levels in plasma samples from the same 137 individuals chronically infected with HIV-1. Median is indicated by black line. Relationship between **B)** p24 and HIV-RNA, **C)** p24 and HIV-DNA and **D)** p24 and CD4+ T cell counts in plasma samples from individuals chronically infected with HIV-1. Correlations were calculated using a nonparametric Spearman test.

Next we aimed to analyze p24 values after cART treatment. The participants at the ANRS REFLATE trial were co-infected with tuberculosis. They initiated antiretroviral treatment (raltegravir (400mg), raltegravir (800mg) or efavirenz (600mg), with tenofovir and lamivudine) 2-8 weeks after treatment for TB was initiated. We compared the _us_p24 values prior to cART initiation to that 24 and 48 weeks after cART initiation in 108 participants. There was a dramatic decrease of p24 levels at weeks 24 and 48 after cART initiation, in line with the decrease in RNA viral load levels observed at the same time points (Figure 5A and (28)). The _us_p24 became undetectable for 89/108 participants after 48 weeks on cART (Figure 5A). In the 19 individuals with _us_p24 above the cutoff at W48, most of them (14/19) had undetectable viral RNA loads at this time point. We observed several differences when we compared the characteristics before cART initiation of these 19 participants with the characteristics of the other participants whose p24 leves were detectable before cART and dropped to undetectable levels with treatment (n=59). Although both subgroups of participants had similar RNA viral loads and _us_p24 levels before treatment, we found a tendency for a lower HIV-RNA/_us_p24 ratio in the participants who maintained detectable _us_p24 after 48 weeks of treatment (Figure 5B). Moreover, these participants had, before treatment, lower cell-associated HIV-DNA levels (Figure 5C) and higher CD4+ T cell counts (Figure 5D) than the participants with undetectable _us_p24 levels after cART. No differences were observed for the same parameters at W48.

**Figure 5.**
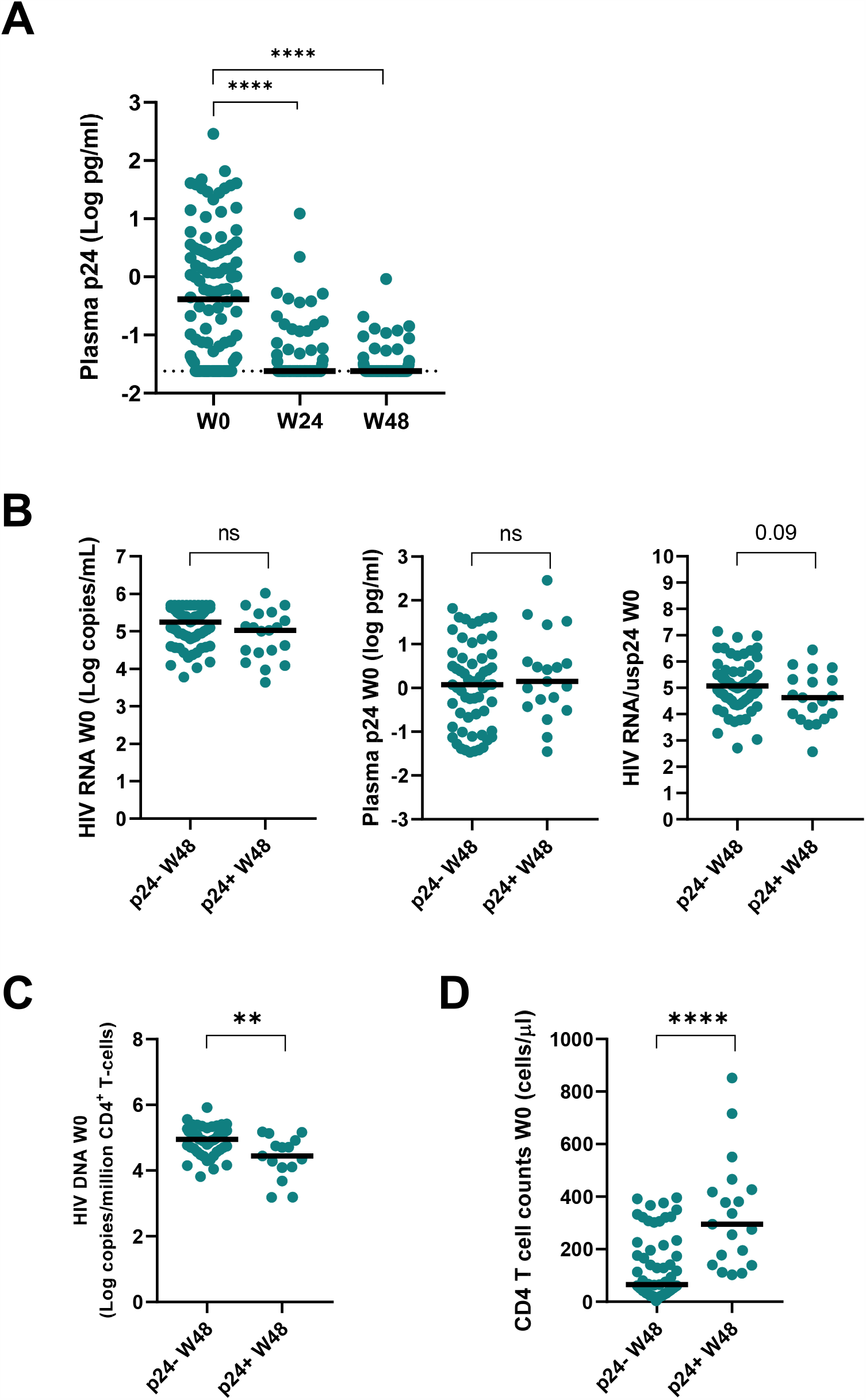
p24 levels in patients chronicaly infected with HIV prior to and after cART initiation. **A)** p24 levels in plasma samples from 108 individuals chronically infected with HIV-1 (ANRS 12180 Reflate TB trial) prior to antiretroviral treatment initiation (W0) and longitudinally monitored at weeks 24 and 48 after cART. Baseline (W0) differences between patients who presented detectable or undetectable p24 levels 48 weeks after cART initiation: **B)** HIV-RNA levels (left panel), p24 (middle panel) and the ratio HIV-RNA/p24 (right panel), **C)** HIV-DNA, **D)** CD4+ T cell counts. Median is indicated by black lines. ** <0.01, **** < 0.0001.

We analyzed the association of _us_p24 levels with other plasma markers measured in the same samples. At W0, _us_p24 levels were positively correlated with D-dimers (r=0.24, p=0.006) and IFNα (r=0.38, p<0.001)(Figure 6A), and these correlations were still statistically significant when we took into consideration only the _us_p24 values above the cutoff (r=0.23, p=0.03; and r=0.25 p=0.02, for D-dimers and IFN-α respectively). We did not find correlations between _us_p24 and CRP, IL-6 or sCD14. We then studied the subgroup of participants who remained detectable for p24 at W48. They also differed for the levels of some inflammatory markers before treatment initiation. While the participants whose _us_p24 levels became undetectable with cART had higher IFNα levels at W0 (Figure 6B), the participants whose _us_p24 levels remained detectable were characterized by higher plasma levels of CRP (Figure 6C), and tended to have higher levels of D-dimers (Figure 6D) and IL-6 (Figure 6E). Altogether, these results show that _us_p24 can be monitored throughout HIV-1 infection and may have a complementary value to other biomarkers. Moreover, the detection of p24 in the plasma of PWH under cART may reveal the presence of a subset of people with particular immunovirological characteristics.

**Figure 6.**
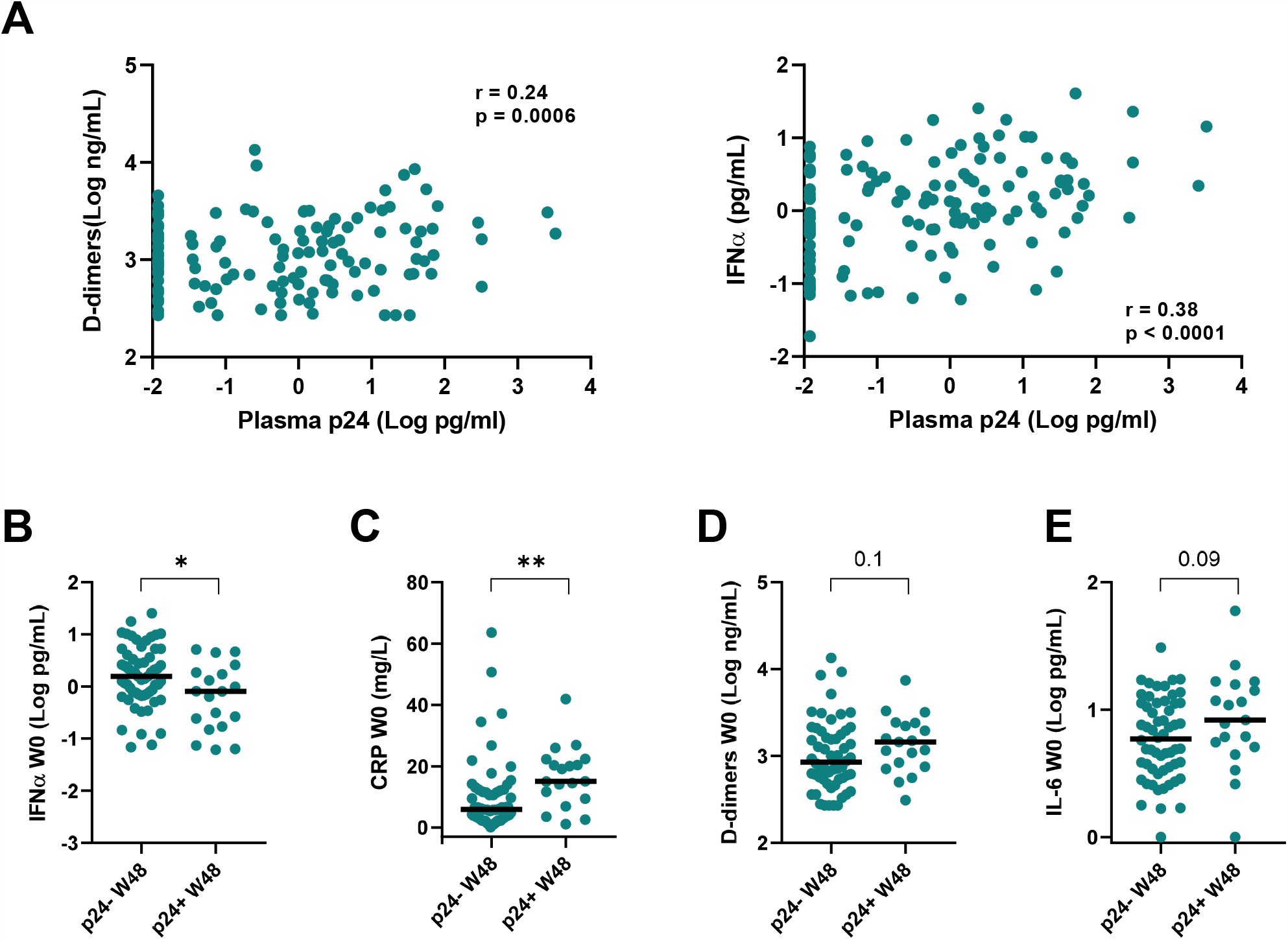
Association of p24 levels and inflammation markers in patients chronicaly infected with HIV prior to and after cART initiation. **A)** Relationship between p24 and D-dimers (Left panel) and IFNα (Right panel) in plasma samples from 137 individuals chronically infected with HIV-1 (ANRS 12180 Reflate TB trial) prior to cART initiation. Correlations were calculated using a nonparametric Spearman test. Baseline (W0) differences between patients who presented detectable or undetectable p24 levels 48 weeks after cART initiation: **B)** IFNα, **C)** CRP, **D)** D-dimers, and **E)** IL-6. Median is indicated by black lines. * <0.05, ** <0.01.

## Discussion

Plasma HIV-RNA and CD4+ T cell counts are commonly used biomarkers to monitor HIV-1 infection. However, these markers do not always reflect the evolution of infection, in particular the occurrence of comorbidities or viral reactivation events during antiretroviral treatment. Here, we developed a method that allowed quantification of _us_p24 despite the presence of anti-p24 antibodies. We show that ultrasensitive p24 quantification is a robust virologic marker associated with HIV-1 RNA and DNA during primary and chronic HIV-1 infection, which recapitulates viral suppression observed upon cART, but may also provide added value regarding events dissociated from viral replication.

In the present work we report a higher frequency of p24 detection in samples with viral loads higher than 10^3^ copies/mL, which correspond to an increment of at least 1 Log when compared to conventional ELISA methods (23, 24, 29). Overall, we estimate that current standard immunoassays would allow p24 detection in a very small fraction (15%) of the samples included in this study. Here, the quantification of p24 was possible in > 70% of tested samples. As compared to nucleic acid quantification, the sensitivity of the _us_p24 assay is still limited, due in part to the amount of plasma that can be tested. The inclusion of a prior ultracentrifugation step represents an increase in the hands-on time, but efficiently increases the sensitivity of immunoassays for quantification of p24 in in plasma samples with RNA viral loads at the magnitude of 10^2^ copies/mL (not shown).

Numerous studies have shown that chronic HIV-related immune activation and inflammation are associated with increased risk of morbidity and mortality in PWH (30-35). Moreover, some plasma markers better predict evolution of HIV-1 infection than viral load or CD4+ T cell counts (36-38). We found that _us_p24 was correlated with plasma levels of the coagulation marker D-dimer during chronic infection. We also found a correlation between _us_p24 and IFNα, which could also be quantified by the ultrasensitive digital assay in the same samples. Some recent studies have shown that markers of HIV-1 transcription may be better associated with systemic inflammation than viremia or the levels of integrated proviruses (39-42). The relative association of available biomarkers of infection with the levels of inflammation may depend on the various factors that sustain chronic inflammation (33, 43).

Strong correlations were found between HIV-RNA and _us_p24 in the samples obtained during primary and chronic infection, and the overall _us_p24 detection rate was similar in the samples from the chronically infected cohort than in those from the PHI cohort (72% vs 78%). However, while in the PHI cohort we could detect p24 in most samples with VL>10^4^ RNA copies/mL (64/68 samples with detectable _us_p24), the _us_p24 detection rate was lower in the chronic samples with equivalent VL (94/121 samples with detectable _us_p24, p=0.008). The relative half-lives of HIV-1 p24 and RNA diverge during the course of infection (44) and the difference of antibodies avidity between PHI and chronic infection could also impact the ICD efficacy (26). It is also important to notice that the samples corresponding to chronic infection came from participants co-infected with HIV-1 and TB, which were characterized by higher inflammation levels than PWH with no TB co-infection, even after 48 weeks of suppressive antiretroviral treatment (28). Additional studies including PWH without co-infections should be done in the future to elucidate whether _us_p24 may in some cases better reflect viral activity in blood and/or in lymphoid tissues than viral RNA. Further studies might also analyze its potential contribution to chronic inflammation. In any case, HIV-1 _us_p24 and RNA levels may provide complementary readouts to monitor infection. It is reasonable to imagine that p24 levels during chronic infection can indicate the presence of viral particles, in particular without cART, but also antigen production by cells carrying non-productive viruses, which frequency increases with duration of infection (45-47). The latter might particularly be the case during antiretroviral treatment. The half-life of p24 outside viral particles was shown to be around 42 days, and thus longer than for viral RNA, which may allow the detection of p24 even after viral replication is suppressed (13). It was indeed intriguing to observe that in some samples p24 remained detectable despite undetectable viral RNA levels. These samples came from participants with marked immunological and virological differences before treatment initiation, although this would need confirmation in a larger group of PWH. It is tempting to speculate nevertheless that these differences might arise from better stimulation of adaptive responses by HIV antigens before treatment initiation. Unfortunately, no analyses of T or B cell activation, nor of HIV-specific responses were available for this study.

In conclusion, this study shows that digital ELISA combined with acid-ICD allows the monitoring of _us_p24 in individuals with primary and chronic HIV-1 infection. This novel tool may provide helpful information to monitor the burden of infection during cART. Moreover, this marker could be helpful to investigate the presence of HIV antigens in samples from participants undergoing interventions aiming at HIV eradication, such as HIV reservoir reactivation in “shock and kill” strategies (48).

## Methods

### Cohorts and ethics statement

Blood samples from HIV-negative controls were obtained from the French Blood Bank (Etablissement Français du Sang) in the context of a collaboration agreement with the Institut Pasteur (C CPSL UNT, number 15/EFS/023). Plasma samples from people with acute and chronic HIV-1 infection monitored for virologic markers at the Hospital Necker-Enfants malades (France) were used to determine the efficacy of ICD and the sensitivity of the Quanterix Simoa p24 assay in plasma samples. Plasma samples from the French National Agency for Research on AIDS and Viral Hepatitis (ANRS) CO6 PRIMO cohort (49) and 12180 Reflate TB trial (50) were used to validate the protocol for ultrasensitive p24 detection combined with ICD. People with primary HIV infection (PHI) were enrolled in the ANRS CO6 PRIMO following the inclusion criteria as described in (49). The cohort was approved by the Ile-de-France-3 Ethics Committee and all patients give their written informed consent. The ANRS 12180 Reflate TB trial was carried out in accordance with the ANRS Ethical Chart for Research in Developing Countries, the Brazilian regulatory requirements for clinical trials and the Declaration of Helsinki. The protocol was approved by national and local ethics committees in Brazil (Comissão Nacional de Ética em Pesquisa [CONEP] and Comitê de Ética em Pesquisa [CEP] at IPEC/FIOCRUZ) and France (Comité de Protection des Personnes de Paris Ile-de-France-I). The experiments were conducted with the understanding and the written informed consent of each participant.

### Samples

EDTA blood samples were collected from each individual analyzed. Plasmas were collected after centrifugation at 1800 rpm for 20 minutes and stored at −80°C.

### Immune-complex dissociation (ICD)

Heat-mediated dissociation (51), with SDS and DTPA (26) and acid dissociation (52-54) methods were tested for compatibility with ultrasensitive HIV-1 Gag p24 detection.

#### Heat-mediated immune-complex dissociation

Plasma samples (100 μL) were diluted 1/3 with distilled water and incubated for 5 minutes at 95°C in a water bath, then cooled to room temperature prior to p24 quantification.

#### Heat-mediated immune-complex dissociation with SDS and DTPA

Plasma samples (100 μL) were diluted 1/3 with a solution of 7mM sodium dodecyl sulfate (SDS) and 1.5 mM diethylenetriaminepentaacetic acid (DTPA), pH 7.2. Samples were incubated for 5 minutes at 95°C in a water bath, then cooled to room temperature prior to p24 quantification.

#### Acid dissociation

Plasma samples (100 μL) were diluted 1:1 with 1.5 M glycine-HCl pH 1.8, incubated 60 minutes at 37°C, and then neutralized with 1.5 M Tris-HCl pH 9.0. In parallel we tested 1.5 M glycine-HCl pH 2.5 and neutralization with 1.5 M Tris-HCl pH 7.5.

### Ultrasensitive p24 (_us_p24) digital immunoassay

Plasma concentration of HIV-1 Gag p24 was determined on a Simoa HD-1 analyzer using the Simoa HIV p24 kit (Quanterix, USA) following manufacturer’s instructions. Plasma samples were thawed at room temperature, centrifuged at 3000rpm for 5 minutes, and then inactivated with Triton X-100 (final concentration 2%) prior to p24 quantification. Four-parameter logistic (4PL) regression fitting was used to estimate the concentration of p24. Samples below the limit of quantification were given an arbitrary value of 0.024 pg/mL based on the established cutoff (calculated as 2.5 standard deviations from the mean of p24 signal in plasma from HIV negative donors).

### IFNα ultrasensitive digital immunoassay

Plasma samples from the ANRS CO6 PRIMO cohort and the ANRS Reflate TB trial were assayed for IFNα. Plasma samples were thawed at room temperature, centrifuged at 3000rpm for 5 minutes, and then inactivated with Triton X-100 (final concentration 2%) prior to IFNα quantification. Plasma concentration of IFNα was determined on a HD-1 analyzer using the Simoa human IFNα kit (Quanterix, USA) following manufacturer’s instructions. Four-parameter logistic (4PL) regression fitting was used to estimate the concentration of IFNα.

### Clinical parameters and inflammation biomarkers

ANRS PRIMO cohort viral RNA loads in plasma were determined with the COBAS AmpliPrep/COBAS TaqMan HIV-1 Test, v2.0 (Roche Diagnostics, Germany). Total cell-associated HIV-1 DNA was quantified with the Generic HIV-1 DNA Cell kit (Biocentric, Bandol, France). CD4+ T-cell counts were determined by flow cytometry as previously described (49, 55). The date of infection was estimated based on the date of symptom onset minus 15 days, or, in asymptomatic patients, the date of the incomplete Western blot finding minus 1 month or the midpoint between a negative and a positive ELISA result (56).

The clinical parameters and inflammation biomarkers for the participants to the ANRS Reflate TB trial were determined as described in (50) and (28). Briefly, CD4+ T-cell counts were determined by flow cytometry. Plasma HIV-RNA was determined either with the COBAS AmpliPrep/COBAS TaqMan HIV-1 Test, v2.0 (Roche Diagnostics, Germany) or the VERSANT HIV-RNA 3.0 assay (bDNA; Bayer, USA). Total cell-associated HIV-1 DNA was quantified with the Generic HIV-1 DNA Cell kit (Biocentric, Bandol, France). hsCRP and IL-6 levels were measured with the high-sensitivity Tina-quant C-Reactive Protein Gen.3 and the Elecsys IL-6 Immunoassay kits, respectively. D-Dimers were quantified using the STA-Liatest D-Di Plus. Human sCD14 was measured with the hCD14 Quantikine ELISA Kit.

### Statistical analyses

Statistical analyses were performed using GraphPad Prism, version 8.3.1. Comparisons between groups were based on the nonparametric Mann-Whitney test. Correlations were performed using the Spearman test. Differences were considered significant at a *P* value <0.05.

## Data Availability

All data is available in the manuscript or upon request. No datasets were generated

## Acknowledgements

This study was funded by the ANRS, the French National Agency of research on AIDS and Viral Hepatitis and by MSDAvenir to A.S.-C. This study was developed in the context of the ANRS RHIVIERA program.

## DECLARATION OF INTERESTS

The authors declare no competing interests.

## Notes

### Competing Interest Statement

The authors have declared no competing interest.

### Author Declarations

Blood samples from HIV‐negative controls were obtained from the French Blood Bank (Etablissement Francais du Sang) in the context of a collaboration agreement with the Institut Pasteur (C CPSL UNT, number 15/EFS/023). Plasma samples from people with acute and chronic HIV‐1 infection monitored for virologic markers at the Hospital Necker‐Enfants malades (France) were used to determine the efficacy of ICD and the sensitivity of the Quanterix Simoa p24 assay in plasma samples. Plasma samples from the French National Agency for Research on AIDS and Viral Hepatitis (ANRS) CO6 PRIMO cohort (NCT03148964) and ANRS 12180 Reflate TB trial (NCT02273765) were used to validate the protocol for ultrasensitive p24 detection combined with ICD. ANRS PRIMO cohort was approved by the Ile‐de‐France‐3 Ethics Committee and all patients give their written informed consent. The ANRS 12180 Reflate TB trial was carried out in accordance with the ANRS Ethical Chart for Research in Developing Countries, the Brazilian regulatory requirements for clinical trials and the Declaration of Helsinki. The protocol was approved by national and local ethics committees in Brazil (Comissao Nacional de Etica em Pesquisa [CONEP] and Comite de Etica em Pesquisa [CEP] at IPEC/FIOCRUZ) and France (Comite de Protection des Personnes de Paris Ile‐de‐France‐I). The experiments were conducted with the understanding and the written informed consent of each participant

